# Publication Bias in Abstracts Presented at the American Diabetes Association Scientific Sessions: A Retrospective Cohort Study

**DOI:** 10.64898/2026.08.26.26361486

**Authors:** Isabel Pinedo-Torres, Alvaro Taype-Rondan, Adai A. Vera-Luza, Paolo A. Zegarra-Lizana, José Luis Rojas-Vilca, Marlon Yovera-Aldana

## Abstract

**Objective:** To determine the publication rate of abstracts presented at the American Diabetes Association Scientific Sessions and to evaluate the association between statistical significance of study results and subsequent publication.

**Research Design and Methods:** We conducted a retrospective cohort study of abstracts presented at the 2018 American Diabetes Association Scientific Sessions. The primary exposure was study result category (statistically significant vs. non-statistically significant findings), and the primary outcome was publication in an indexed journal within 5 years after conference presentation. Publication status was determined through PubMed/MEDLINE and Scopus searches. Adjusted relative risks (RRs) and 95% CIs were estimated using generalized linear models with Poisson distribution and robust variance.

**Results:** Among 541 included abstracts, 321 (59.3%) were subsequently published in indexed journals. Abstracts reporting statistically significant findings had a higher publication rate than those reporting non-statistically significant findings (61.9% vs. 42.3%; p=0.002). In the adjusted analysis, abstracts with non-statistically significant findings had a lower likelihood of publication compared with those reporting statistically significant findings (adjusted RR 0.71 [95% CI 0.54–0.93]; p=0.012).

**Conclusions:** Approximately four in ten abstracts presented at the ADA Scientific Sessions were not published within 5 years. Abstracts reporting non-statistically significant findings had a lower likelihood of subsequent publication, suggesting persistent publication bias in diabetology research. Future initiatives promoting the interpretation of effect estimates, confidence intervals and clinical relevance, rather than statistical significance alone, may help reduce selective dissemination of evidence

## Introduction

Diabetes mellitus is one of the leading global public health challenges, affecting an estimated 537 million adults worldwide, with projections reaching 783 million by 2045 [1,2]. This increasing burden has been accompanied by substantial growth in diabetes research, generating a large volume of scientific evidence intended to improve prevention, diagnosis, and treatment [3]. Scientific meetings, particularly the American Diabetes Association (ADA) Scientific Sessions, play a central role in the early dissemination of this research and provide an opportunity for rapid communication of new findings before full publication [4]. However, presenting an abstract at these meetings does not guarantee subsequent publication in a peer-reviewed journal, which remains the primary mechanism for ensuring the validation and accessibility of evidence.

Previous studies indicate that publication rates of conference abstracts vary considerably across medical specialties. A Cochrane review determined that fewer than half of the studies presented as abstracts are published as full articles within a 10-year period [5]. Specifically in the field of endocrinology, a conversion rate from abstract to full article of approximately 31.7% has been reported [6]. Various factors influence this probability, including study design (randomized trials have higher success rates), larger sample sizes (n≥100), multicenter collaboration, and oral versus poster presentation [7].

Among these factors, the statistical significance of study findings appears to play an important role in subsequent publication. Previous evidence suggests that studies reporting statistically significant results are more likely to be published and cited than those with non-statistically significant findings [8]. This selective dissemination may contribute to an overrepresentation of statistically significant findings in the biomedical literature. Furthermore, evidence suggests that some specialized journals maintain an implicit preference for statistically significant outcomes, which may discourage dissemination of confirmatory, neutral, or safety-related studies [9]. Consequently, publication bias may distort the available evidence base used in clinical decision-making and evidence synthesis [10].

Although publication bias has been documented across several medical specialties, evidence remains limited for diabetology and, in particular, for abstracts presented at the ADA Scientific Sessions. Evaluating the transition from conference presentation to full-text publication provides an opportunity to assess potential selective dissemination of research findings within this field. Therefore, this study aimed to determine the publication rate of abstracts presented at the 2018 American Diabetes Association Scientific Sessions and to assess the association between the statistical significance of study findings and subsequent publication in indexed journals over a 5-year follow-up period.

## Methods

### Study Design and Data Source

We conducted a retrospective cohort study of abstracts presented at the 2018 Scientific Sessions of the American Diabetes Association [11]. Individual conference abstracts were considered the unit of analysis and were followed for five years to determine subsequent publication in indexed scientific journals. A five-year follow-up period was selected to allow sufficient time for subsequent publication, as most conference abstracts that ultimately achieve publication do so within this timeframe [9]. This study was reported according to the Strengthening the Reporting of Observational Studies in Epidemiology guideline **(S1 table)** [12].

### Population and Eligibility Criteria

Eligible abstracts included primary and secondary analytical studies in any field related to diabetes and its complications, including both human and animal research conducted worldwide. We excluded abstracts unavailable on the ADA website, diagnostic accuracy studies, qualitative studies, descriptive studies without hypothesis testing that precluded classification according to the statistical significance of the primary study findings, studies in which the p-value for the primary outcome was not reported, studies with fewer than 10 participants, and studies already published before conference presentation.

### Sampling and Sample Size

The sampling frame consisted of the digital abstract book from the 2018 ADA Scientific Sessions. Sample size was estimated using OpenEpi version 3.1 based on a previously reported publication rate of 36.6% for conference abstracts, with a 95% confidence level, resulting in a minimum required sample of 354 abstracts [13]. To preserve statistical power after applying the eligibility criteria, a larger random sample was initially selected. A simple random sampling strategy was implemented using EpiData version 4, ensuring that each abstract had an equal probability of selection. A total of 541 abstracts met the eligibility criteria and were included in the final analysis.

### Exposure, outcome and other variables

The primary exposure was the statistical significance of study findings (statistically significant vs. non-statistically significant). Studies were classified as reporting statistically significant findings when the primary hypothesis test yielded a p-value ≤0.05 and as reporting non-statistically significant findings when the p-value was >0.05.

The primary outcome was publication in an indexed journal within five years after conference presentation. Publication status was determined through searches in PubMed/MEDLINE and Scopus using the first author’s name and keywords from the abstract title. Potential matches were manually verified by comparing the research question and primary objective between the abstract and the published article. When multiple publications corresponded to the same abstract, the earliest full-text publication was considered. Abstracts without an identifiable publication were classified as unpublished.

Additional study-level variables included session type (general session, moderated session, oral presentation, or online-only), thematic category, study design (basic/preclinical, observational, experimental, or evidence synthesis/bibliometric), sample size, and number of participating centers. Research team characteristics included number of authors and disclosure of conflicts of interest. Presenter characteristics included institutional affiliation, continent of affiliation, primary language, and research experience. Presenter research experience was assessed using the number of publications attributed to the presenting author in ORCID and Google Scholar. Author identity was verified by cross-checking names, institutional affiliations, research topics, and publicly available academic profiles. Primary language was inferred from the country of institutional affiliation and publicly available academic profiles of the presenting author and should therefore be interpreted as a proxy measure.

Among published studies, secondary outcomes included time to publication, journal region, publication model (diamond, gold, or hybrid open access), and journal impact metrics, including SCImago Journal Rank (SJR), quartile ranking, and H-index. Diamond open-access journals were defined as those without article processing charges (APCs), gold open-access journals as fully open-access journals requiring an APC, and hybrid journals as subscription-based journals offering an open-access publication option.

### Data collection

Conference abstracts were retrieved from the official abstract book of the 2018 ADA Scientific Sessions (https://plan.core-apps.com/tristar_ada18/abstracts). Publication searches were conducted between January and March 2024. Data extraction and publication searches were independently performed by A.A.V.-L. and P.A.Z.-L. Cases in which publication status or article matching remained uncertain were reviewed by I.P.-T. and resolved through discussion with the research team until consensus was reached. Missing data were handled using available-case analysis for descriptive and bivariate analyses and complete-case analysis for the multivariable model. No data were imputed.

### Statistical Analysis

Statistical analyses were performed using Stata version 16. Continuous variables were summarized as mean ± standard deviation or median with interquartile range (IQR), according to their distribution, whereas categorical variables were reported as frequencies and percentages.

Bivariate analyses were conducted using the chi-square test, Fisher’s exact test, or Mann– Whitney U test, as appropriate. To evaluate the association between statistical significance of study findings and publication, generalized linear models with Poisson distribution, log link, and robust variance were used to estimate relative risks (RRs) and 95% confidence intervals (CIs) [14]. This approach was selected to directly estimate RRs because publication was a common outcome, for which odds ratios may differ substantially from RRs [15].

Confounder selection for the adjusted model was guided by a directed acyclic graph (Supplementary figure 1) and theoretical relevance. Study design, participating centers, number of authors, conflicts of interest, and presenting author research experience were considered potential confounders because they could influence both study result type and subsequent publication probability. Google Scholar-indexed publications were used as the measure of presenting author research experience in the adjusted model because this source provided broader publication coverage than ORCID. Because the number of Google Scholar-indexed publications showed evidence of non-linearity on its original scale, it was transformed as ln(x+1) before inclusion in the multivariable model. Variables considered downstream of the exposure–outcome relationship were not included in the adjusted model to avoid overadjustment [16].

Model convergence and stability of estimates were assessed by evaluating convergence diagnostics and consistency between crude and adjusted estimates. Multicollinearity among covariates included in the adjusted model was assessed using variance inflation factors, and no evidence of problematic collinearity was identified. A two-sided p-value <0.05 was considered statistically significant.

### Ethical Considerations

This study analyzed publicly available conference abstracts and did not involve human participants, identifiable personal information, or patient-level data. Because the unit of analysis was published conference abstracts rather than human participants, institutional ethics committee review was not required under local regulations.

## Results

### Abstracts Selection

Of the 2,665 abstracts presented during the 2018 Scientific Sessions of the American Diabetes Association, 708 were randomly selected. After applying the eligibility criteria, 541 abstracts were included in the final analysis **(Figure 1).**

**Figure 1.**
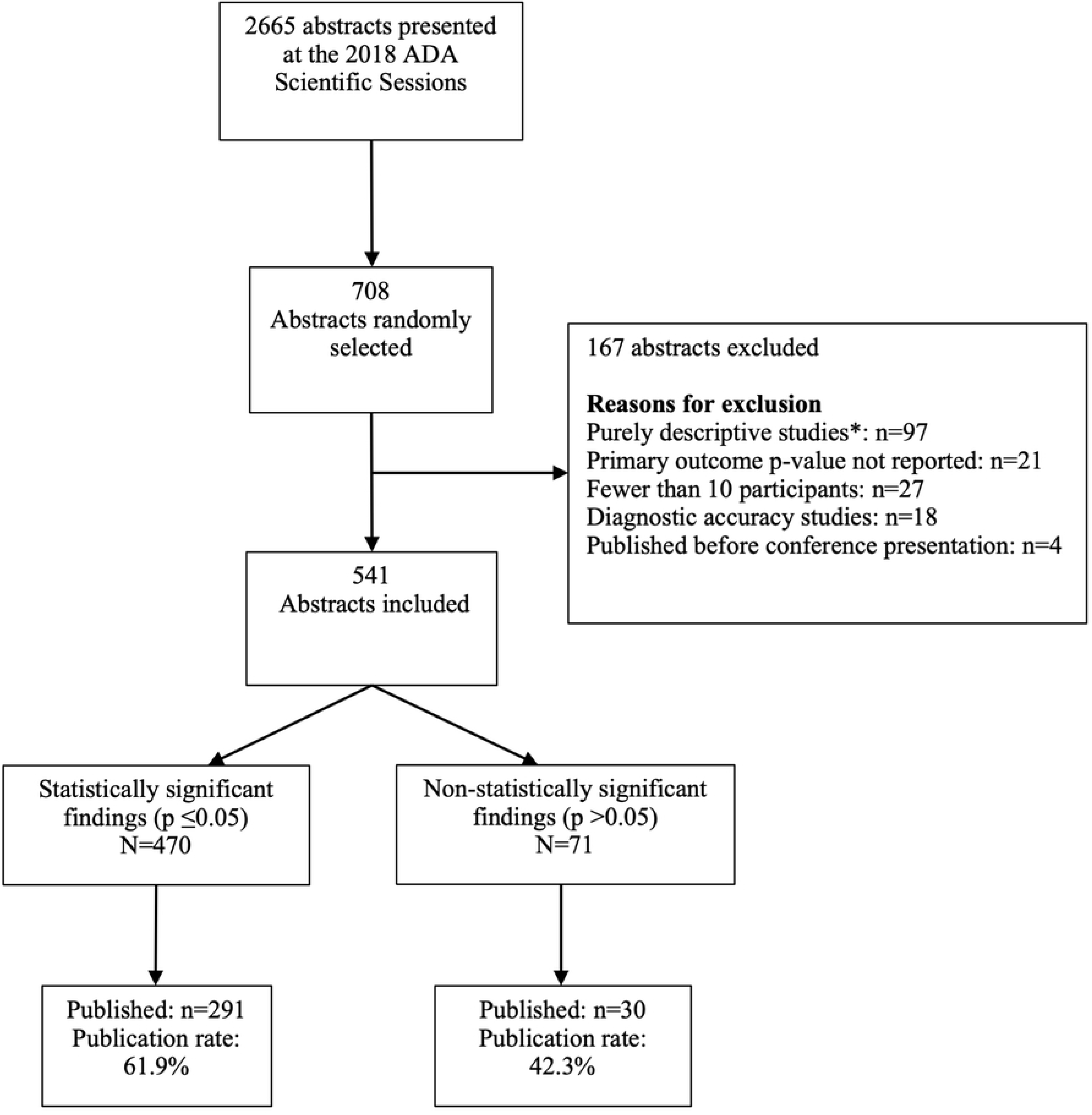
Flowchart of abstract selection. * Studies without analytical hypothesis testing. Abbreviations: ADA, American Diabetes Association; TP, publication rate.

### General Characteristics

Among the 541 included abstracts, 59.3% (n=321) were subsequently published in indexed journals within a five-year follow-up period. Non-statistically significant findings were identified in 13.1% (n=71) of the studies. The most common study design was observational (62.5%), followed by experimental studies (25.9%). The median number of authors was six, and 60.6% of presenting authors had English as their primary language.

Among published abstracts, 59.2% (n=190) had an observational design, whereas only 9.4% (n=30) reported non-statistically significant findings. The median number of authors among published studies was seven, and 59.2% of presenting authors had English as their primary language **(Table 1).**

**Table 1.** Characteristics of abstracts presented at the 2018 Scientific Sessions of the American Diabetes Association according to publication status after 5 years of follow-up.

| Variable | Included abstracts<br>(n=541) n (%) | Published abstracts within<br>5 years (n=321) n (%) |
| --- | --- | --- |
| **Study characteristics** |  |  |
| **Study design** |  |  |
| Basic science / preclinical | 53 (9.8) | 25 (7.8) |
| Observational | 338 (62.5) | 190 (59.2) |
| Experimental | 140 (25.9) | 103 (32.1) |
| Bibliometric / systematic review | 10 (1.9) | 3 (0.9) |
| **Sample size,** |  |  |
| Median [IQR] | 202 [46–1173] | 196.5 [47–1164.5] |
| **Participating centers** |  |  |
| Single-center | 383 (70.8) | 223 (69.5) |
| Multicenter | 158 (29.2) | 98 (30.5) |
| **Statistical significance of findings\*** |  |  |
| Statistically significant | 470 (86.9) | 291 (90.7) |
| Non-statistically significant | 71 (13.1) | 30 (9.4) |
| **Time to publication (months)\*\*** |  |  |
| Median [IQR] | — | 16 [8–28] |
| **Research team characteristics** |  |  |
| **Number of authors** |  |  |
| Median [IQR] | 6 [4–9] | 7 [5–9] |
| **Conflicts of interest** |  |  |
| Absent | 248 (45.8) | 145 (45.2) |
| Present | 293 (54.2) | 176 (54.8) |
| **Presenting author characteristics** |  |  |
| **Continent of affiliation** |  |  |
| North America | 292 (54.8) | 172 (53.6) |
| South America | 11 (2.1) | 6 (1.9) |
| Europe | 120 (22.5) | 74 (23.1) |
| Asia | 103 (19.3) | 65 (20.3) |
| Africa | 1 (0.2) | 0 |
| Oceania | 6 (1.1) | 4 (1.3) |
| **Primary language** |  |  |
| English | 323 (60.6) | 190 (59.2) |
| French | 23 (4.3) | 16 (5.0) |
| Spanish | 10 (1.9) | 4 (1.3) |
| Portuguese | 8 (1.5) | 6 (1.9) |
| Mandarin Chinese | 32 (6.0) | 22 (6.9) |
| Other | 137 (25.7) | 83 (25.9) |
| **Research experience** |  |  |
| ORCID-indexed publications, Median [IQR] | 12 [2–50] | 14 [5–60] |
| Google Scholar-indexed publications, Median [IQR] | 86 [34–233] | 108 [42–283] |

ADA, American Diabetes Association; IQR, interquartile range; ORCID, Open Researcher and Contributor ID. \* Non-statistically significant findings were defined as studies in which the primary hypothesis test yielded a p-value >0.05. \*\* Time to publication was calculated from the date of conference presentation (June 2018) to publication in an indexed journal.

### Characteristics of the Journals in Which the Abstracts Were Published

Among the published abstracts, studies presented in moderated sessions had the highest median SCImago Journal Rank (SJR) (median: 1.90; IQR: 1.11–2.62), whereas abstracts presented orally had the highest median H-index (median: 166; IQR: 120–391). A total of 261 abstracts were published in Q1 journals **(S2 Table).** Regarding journal origin and publication model, 145 abstracts were published in North American journals and 155 in European journals. Most publications appeared in gold open-access journals (n=209), whereas only one article was published in a diamond open-access journal without article processing charges **(S3 Table)**.

### Association Between Study Characteristics and Publication

Among studies reporting statistically significant findings, 61.9% (n=291) were subsequently published, compared with 42.3% (n=30) among studies with non-statistically significant findings (p=0.002). Other variables significantly associated with publication included study design (p<0.001), number of authors (p=0.014), and presenting author research experience, measured by the number of publications indexed in ORCID (p=0.012) and Google Scholar (p<0.001) **(Table 2).**

**Table 2.** Association between study, research team, and presenting author characteristics and subsequent publication of abstracts presented at the 2018 Scientific Sessions of the American Diabetes Association (n=541).

| Variable | Published n (%) | Not published n (%) | p-value |
| --- | --- | --- | --- |
| **Study characteristics** |  |  |  |
| **Study design** |  |  |  |
| Basic science / preclinical | 25 (47.2) | 28 (52.8) | <0.001<sup>a</sup> |
| Observational | 190 (56.2) | 148 (43.8) |  |
| Experimental | 103 (73.6) | 37 (26.4) |  |
| Evidence synthesis/bibliometric | 3 (30.0) | 7 (70.0) |  |
| **Sample size, median [IQR]** | 196 [47–1164.5] | 206 [46–1177] | 0.853<sup>b</sup> |
| **Participating centers** |  |  |  |
| Single-center | 223 (58.2) | 160 (41.8) | 0.413<sup>a</sup> |
| Multicenter | 98 (62.0) | 60 (38.0) |  |
| **Statistical significance of findings\*** |  |  |  |
| Statistically significant | 291 (61.9) | 179 (38.1) | 0.002<sup>a</sup> |
| Non-statistically significant | 30 (42.3) | 41 (57.7) |  |
| **Research team characteristics** |  |  |  |
| **Number of authors, median [IQR]** | 7 [5–9] | 6 [4–8] | 0.014<sup>b</sup> |
| **Conflicts of interest** |  |  |  |
| Absent | 145 (58.5) | 103 (41.5) | 0.706<sup>a</sup> |
| Present | 176 (60.1) | 117 (39.9) |  |
| **Presenting author characteristics** |  |  |  |
| **Continent of affiliation** |  |  |  |
| North America | 172 (58.9) | 120 (41.1) | 0.839<sup>c</sup> |
| South America | 6 (54.5) | 5 (45.5) |  |
| Europe | 74 (61.7) | 46 (38.3) |  |
| Asia | 65 (63.1) | 38 (36.9) |  |
| Africa | 0 | 1 (100) |  |
| Oceania | 4 (66.7) | 2 (33.3) |  |
| **Primary language** |  |  |  |
| English | 190 (58.8) | 133 (41.2) | 0.478<sup>a</sup> |
| French | 16 (69.6) | 7 (30.4) |  |
| Spanish | 4 (40.0) | 6 (60.0) |  |
| Portuguese | 6 (75.0) | 2 (25.0) |  |
| Mandarin Chinese | 22 (68.8) | 10 (31.2) |  |
| Other | 83 (60.6) | 54 (39.4) |  |
| **Research experience** |  |  |  |
| ORCID-indexed publications,<br>median [IQR] | 14 [5–60] | 8 [1–42] | 0.012<sup>b</sup> |
| Google Scholar-indexed<br>publications, median [IQR]<br>(n=519) | 108 [42–283] | 63 [23–178] | <0.001<sup>b</sup> |

ADA, American Diabetes Association; IQR, interquartile range; ORCID, Open Researcher and Contributor ID. Values are presented as n (%) unless otherwise indicated. The total study sample comprised 541 abstracts; denominators may vary because of missing data for some presenting author characteristics. \*Non-statistically significant findings were defined as studies in which the primary hypothesis test yielded a p-value >0.05. <sup>a</sup>p-values obtained using the chi-square test. <sup>b</sup>p-values obtained using the Mann– Whitney U test. <sup>c</sup>p-values obtained using Fisher’s exact test.

### Association Between Statistical Significance of Findings and Publication

Compared with abstracts reporting statistically significant findings, abstracts with non-statistically significant findings had a 29% lower likelihood of publication in indexed journals within five years after presentation at the 2018 ADA Scientific Sessions. The adjusted analysis included 519 abstracts with complete data for all model covariates. After adjustment for study design, number of participating centers, number of authors, conflicts of interest, and presenting author research experience (adjusted RR: 0.71; 95% CI: 0.54– 0.93; p=0.012) **(Table 3).**

**Table 3.**
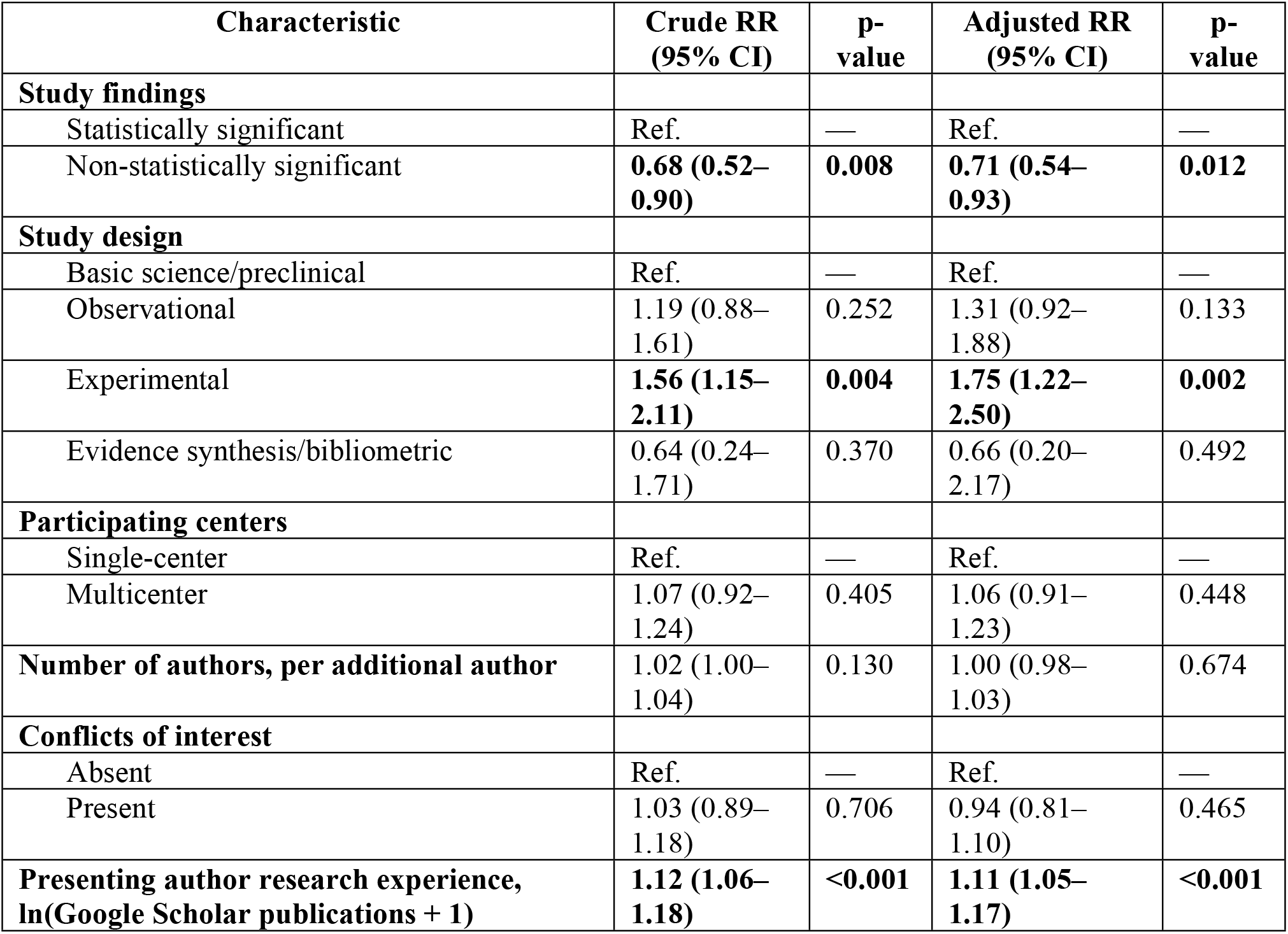
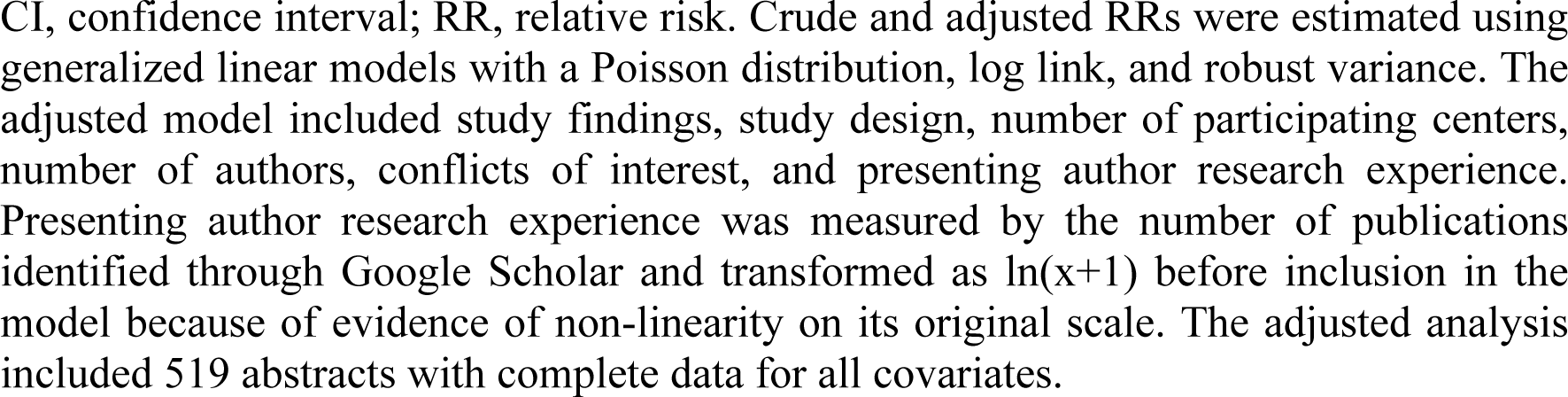
Association between study characteristics and subsequent publication of abstracts presented at the 2018 Scientific Sessions of the American Diabetes Association.

| Characteristic | Crude RR (95% CI) | p-value | Adjusted RR (95% CI) | p-value |
| --- | --- | --- | --- | --- |
| **Study findings** |  |  |  |  |
| Statistically significant | Ref. | — | Ref. | — |
| Non-statistically significant | **0.68 (0.52–0.90)** | **0.008** | **0.71 (0.54–0.93)** | **0.012** |
| **Study design** |  |  |  |  |
| Basic science/preclinical | Ref. | — | Ref. | — |
| Observational | 1.19 (0.88–1.61) | 0.252 | 1.31 (0.92–1.88) | 0.133 |
| Experimental | **1.56 (1.15–2.11)** | **0.004** | **1.75 (1.22–2.50)** | **0.002** |
| Evidence synthesis/bibliometric | 0.64 (0.24–1.71) | 0.370 | 0.66 (0.20–2.17) | 0.492 |
| **Participating centers** |  |  |  |  |
| Single-center | Ref. | — | Ref. | — |
| Multicenter | 1.07 (0.92–1.24) | 0.405 | 1.06 (0.91–1.23) | 0.448 |
| **Number of authors, per additional author** | 1.02 (1.00–1.04) | 0.130 | 1.00 (0.98–1.03) | 0.674 |
| **Conflicts of interest** |  |  |  |  |
| Absent | Ref. | — | Ref. | — |
| Present | 1.03 (0.89–1.18) | 0.706 | 0.94 (0.81–1.10) | 0.465 |
| **Presenting author research experience, ln(Google Scholar publications + 1)** | **1.12 (1.06–1.18)** | **<0.001** | **1.11 (1.05–1.17)** | **<0.001** |

CI, confidence interval; RR, relative risk. Crude and adjusted RRs were estimated using generalized linear models with a Poisson distribution, log link, and robust variance. The adjusted model included study findings, study design, number of participating centers, number of authors, conflicts of interest, and presenting author research experience. Presenting author research experience was measured by the number of publications identified through Google Scholar and transformed as $\ln(x+1)$ before inclusion in the model because of evidence of non-linearity on its original scale. The adjusted analysis included 519 abstracts with complete data for all covariates.

## Discussion

### Principal findings

In this retrospective cohort study of abstracts presented at the 2018 Scientific Sessions of the American Diabetes Association, abstracts reporting non-statistically significant findings had a significantly lower probability of subsequent publication in indexed journals, even after adjustment for study design, participating centers, number of authors, conflicts of interest, and presenting author research experience. In addition, publication was associated with study design and greater presenting author research experience.

### Comparison with other studies

The publication rate identified in our study was higher than that reported in several previous studies of conference abstracts, although publication rates vary considerably across medical specialties. In endocrinology and endocrine surgery, a conversion rate of approximately 31.7% has been reported [6], whereas in the Society of General Internal Medicine, nearly 47% of abstracts reach full publication, with clinical trials and multicenter studies showing the highest likelihood of success [17].

When comparing with other specialties, publication bias favoring statistically significant findings appears to be a broader phenomenon in biomedical research. In ophthalmology, studies with statistically significant results have been reported to be published more frequently and to appear in journals with higher impact factors [18]. Similarly, in orthopedic surgery, publication rates have been reported to differ between oral presentations and posters, suggesting that presentation format may be associated with subsequent publication [19]. In upper gastrointestinal surgery, approximately 32% of conference abstracts have subsequently reached full publication [20].

The five-year follow-up period used in our study allowed sufficient time to capture delayed publication. Systematic reviews examining the publication fate of biomedical conference abstracts indicate that, although many studies are published within the first two years after presentation, additional publications continue to occur during longer follow-up periods [11]. Compared with studies in geriatrics, where publication rates of 24.8% have been reported over similar follow-up periods, the publication rate observed among ADA abstracts was higher. However, comparisons across specialties should be interpreted cautiously because conference characteristics, study designs, publication practices, and methods used to ascertain publication may differ between studies [7].

### Interpretation and implications

Several factors may contribute to the lower publication probability of studies reporting non-statistically significant findings. Selective dissemination may occur at different stages of the research dissemination process. At the investigator level, studies with non-statistically significant or less favorable findings may be perceived as less novel or less likely to be accepted, potentially reducing the likelihood of manuscript preparation or submission. Lack of time, competing research priorities, and decisions not to pursue publication have also been reported as important reasons why conference abstracts do not progress to full-text publication [21]. Editorial and peer-review processes may also contribute to selective dissemination if perceived novelty, clinical relevance, or the direction and statistical significance of findings influence publication decisions [21,22]. Another potential mechanism is time-lag bias, whereby studies with less favorable or non-statistically significant findings may take longer to reach publication [22]. Although our study did not assess investigators’ reasons for non-publication, editorial decision-making, or differences in time to publication according to statistical significance, these mechanisms may individually or collectively contribute to the lower publication probability observed among abstracts with non-statistically significant findings during the five-year follow-up period.

This selective dissemination may contribute to an imbalance in the scientific literature, with statistically significant findings being overrepresented and non-statistically significant findings being comparatively underrepresented. Current methodological recommendations emphasize interpreting effect estimates and their confidence intervals rather than relying on statistical significance alone, particularly when assessing clinical relevance, safety outcomes, and equivalence or non-inferiority [23]. In this context, scientific societies and journals should encourage the dissemination of methodologically rigorous studies regardless of the statistical significance of their findings, thereby promoting a more complete and balanced evidence base [24].

Future studies should evaluate publication bias across multiple years, medical specialties, and scientific conferences to determine whether the observed association is consistent across research settings. Studies examining time to publication according to the direction and statistical significance of findings could help characterize the contribution of time-lag bias. In addition, distinguishing between non-submission by investigators, rejection during peer review, and other reasons for non-publication would help identify the stages of the dissemination process at which selective publication is most likely to occur.

### Strengths and limitations

This study also has several strengths. We used a systematic publication-tracking strategy combining PubMed/MEDLINE and Scopus searches, together with manual verification of author names, study objectives, and research characteristics to reduce the risk of publication misclassification [11]. The five-year follow-up period allowed sufficient time to identify delayed publications. In addition, robust Poisson regression provided direct estimates of relative risks, and a directed acyclic graph was used to guide confounder selection [25]. Reporting according to the STROBE guideline further supports transparency and reproducibility [12].

Despite its contributions, this study has several limitations. First, we relied on the completeness of conference abstracts, which often lack detailed methodological data; if titles or authors changed significantly before publication, some works might have been missed, leading to a potential underestimation of the overall conversion rate. Second, our results are primarily applicable to large international congresses like the ADA, and the exclusion of journals not indexed in PubMed or Scopus may also suggest that the true publication rate is slightly higher than reported [26]. Third, classification of study findings using a binary p-value threshold may have introduced misclassification because this approach does not account for exploratory or secondary analyses, equivalence or non-inferiority designs, or the clinical relevance and precision of effect estimates. Some studies classified as having non-statistically significant findings may therefore have provided clinically meaningful or methodologically informative evidence. This classification could have influenced the magnitude of the observed association; consequently, our findings should be interpreted within the context of conventional statistical significance frameworks commonly used in biomedical research [27,28]. Finally, the relatively small number of abstracts reporting non-statistically significant findings may have limited the precision of some estimates and subgroup comparisons.

## Conclusion

This study found that abstracts reporting non-statistically significant findings at the ADA Scientific Sessions had a lower probability of subsequent publication compared with those reporting statistically significant findings. These findings are consistent with the persistence of publication bias in diabetology research. Greater emphasis on clinical relevance, effect sizes, and confidence intervals may help promote a more balanced dissemination of scientific evidence, regardless of statistical significance.

## Data Availability

The de-identified minimal dataset required to replicate the findings of this study is provided as Supporting Information

## Supplementary material

S1 Table. STROBE Statement—checklist of items that should be included in reports of observational studies

S2 Table. Journal impact metrics of journals publishing abstracts presented at the 2018 American Diabetes Association Scientific Sessions (n=321)

S3 Table. Geographic distribution and publication model of journals in which abstracts from the 2018 American Diabetes Association Scientific Sessions were published, according to research area and presentation format

S1 Figure. Directed acyclic graph (DAG) representing the hypothesized relationships between statistical significance of study findings and subsequent publication.

S1 File. Analysis dataset.

